# Linguistic Validation of the Rett Syndrome Behavior Questionnaire Spanish Translation: a Two-Stage Caregiver Study Across Latin America

**DOI:** 10.64898/2026.04.16.26349544

**Authors:** Melanie Polo-Sánchez, Andrea Carolina Lesmes, Neal Muni, Frederic Vigneault, Richard Novak

**Author notes:** Corresponding Author Information: Richard Novak, PhD, Unravel Biosciences, Inc., 196 Boston Ave, Suite 1000, Medford, MA 02155 USA. Funding /Sponsor statement: This study was funded and sponsored by Unravel Biosciences. Ethics statement: The Rett Syndrome Behaviour Questionnaire (RSBQ) study involved voluntary caregiver participation. An Institutional Review Board (IRB) at the University of Massachusetts Boston reviewed and approved this study (IRB ID #3700). Participants were assigned coded identifiers; identifying information was stored securely and removed following successful data collection.

## Abstract

**Background:** Rett Syndrome (RTT) is a severe neurodevelopmental disorder affecting approximately 1 in 10,000 live female births worldwide. The Rett Syndrome Behaviour Questionnaire (RSBQ), remains one of the most widely used standardized behavioral assessment tools for RTT. However, the RSBQ was originally validated only in British English, limiting its applicability for Spanish-speaking caregivers and clinical centers across Latin America and Spain.

**Objective:** The primary aim of this study was to develop and validate the comprehension of the Spanish translation of the RSBQ to ensure cultural and linguistic equivalence, enhance data reliability, and facilitate earlier, more accurate clinical assessments among Spanish-speaking RTT populations.

**Methods:** Surveys were administered in two phases to Spanish-speaking caregivers between November 2023 and September 2025. Phase I consisted of 12 guided survey administrations with participants being able to ask clarifying questions and offer linguistic modifications of RSBQ questions. Phase II consisted of independent online administration of the refined Spanish RSBQ and a retest at least 7 days later. Participants were recruited through direct outreach and supported virtually during questionnaire completion.

**Results:** Following data cleaning and quality control, a total of 51 caregivers successfully completed both surveys. The Spanish RSBQ demonstrated high caregiver comprehension and strong engagement across multiple Latin American countries, including Argentina, Mexico, and Peru. Responses were highly correlated between test and retest timepoints, and no question showed biased response distributions. A slight effect of response interval on test-retest correlation was observed, potentially indicating the impact of natural disease progression confounding retest evaluation for long (>80 day) intervals; however this effect did not impact the overall linguistic validation results as analysis of only <21 day test-retest responders confirmed the findings.

**Conclusions:** This linguistic validation study represents the first formal step toward the clinical validation of the Spanish RSBQ, enabling broader inclusion of Spanish-speaking populations in RTT research. The collaborative, bilingual data collection strategy proved both feasible and effective, paving the way for multinational trials and expanding therapeutic accessibility through localized, patient-centered innovation.

## INTRODUCTION

Rare diseases are broadly defined as conditions that affect a very small fraction of the population. For example, Panama’s rare disease law (one of the first in Central America) defines a rare condition as one affecting fewer than 1 in 2,000 people. Such diseases are often chronic, debilitating, and life-threatening. Rett Syndrome (RTT) is one such rare disorder – a neurodevelopmental condition first described in 1966 by Dr. Andreas Rett – which occurs almost exclusively in girls. Rett Syndrome is caused by mutations in the X-linked MECP2 gene and leads to severe cognitive, motor, and autonomic impairments.^1^ Children with RTT typically develop normally for 6–18 months, then enter a regression phase where they lose acquired skills (like speech and purposeful hand use) and develop hallmark features such as hand stereotypies (e.g. hand-wringing) and gait abnormalities.^2^ The disorder also involves multiple comorbidities – seizures, breathing irregularities, sleep disturbances, and others – that require lifelong care. Importantly, Rett Syndrome is classified as an orphan disease: its global incidence is roughly 1 in every 10,000 female births. This translates to an estimated prevalence on the order of 5–10 per 100,000 girls and women worldwide. With such low prevalence, RTT meets the criterion of a rare disease in every country, presenting unique challenges for diagnosis, research, and treatment development.

### Rising Awareness and Diagnoses in Latin America

Historically, Rett Syndrome has been underdiagnosed in regions like Latin America, in part due to limited access to genetic testing and low awareness. Until recently, data on RTT in Latin America were scarce – for example, the first documented cases in some countries appeared only in the last two decades. However, in the past five years there has been growing recognition and reporting of RTT cases across Latin America, suggesting that incidence is not so much biologically increasing as it is finally being identified and recorded. Advances in genetic diagnostics and growing clinician awareness have led to more girls being correctly diagnosed with RTT rather than being misclassified under autism or cerebral palsy. For instance, a 2024 pilot program in Chile, prompted by the local Rett patient foundation, implemented genomic testing for clinically suspected RTT cases – confirming MECP2 mutations in approximately 40% of those patients.^3^ This illustrates how greater awareness and better tools are uncovering more RTT cases in the region, many of which would have been missed in the past. In general, the expected incidence of RTT in Latin America should mirror global rates (roughly 1 in 10,000 female births), which implies that for every 10,000 baby girls born, one may have Rett Syndrome. With many Latin American countries experiencing large birth cohorts, there are potentially hundreds of RTT patients region-wide – underscoring the need for local resources to support them.

Crucially, governments in the region have begun to take an interest in rare diseases including RTT. Panama offers a notable example: it passed a landmark rare diseases law in 2014 (updated in 2022) to guarantee social protection for people with rare, low-frequency, and orphan diseases. This law, now recognized as a model in Central America, declares rare diseases a matter of national interest and seeks to improve patients’ access to diagnosis, specialized treatments, and rehabilitation services. The renewed government focus (with the 2022 amendments) is aimed at creating rare disease registries, funding genomic tests, and integrating care for affected individuals. Such policy developments reflect a broader trend across Latin America: rising advocacy and policy support are shining a spotlight on rare diseases, leading to better identification of conditions like RTT. In short, the region is witnessing a positive feedback loop – more awareness and policy support lead to more diagnoses, which in turn highlight the urgent need for localized tools and interventions for Rett Syndrome.

### The Rett Syndrome Behaviour Questionnaire (RSBQ)

One key tool in managing RTT is the Rett Syndrome Behaviour Questionnaire (RSBQ). Developed over 20 years ago (prior to discovery of the MECP2 gene), the RSBQ was the first standardized instrument designed specifically to capture the unique behavioral and emotional features of Rett Syndrome.^4^ It is a caregiver-completed questionnaire comprising 45 items that survey a wide range of RTT symptoms and behaviors. Each item describes a potential symptom (for example, “spells of inconsolable crying for no apparent reason” or “breath holds”) and the caregiver rates how often it has been true for their child in the past two weeks (e.g. “never”, “sometimes”, or “often”). These item scores (typically 0, 1, or 2 points) are summed to produce a total score, as well as scores for 8 subscale domains that reflect core features of RTT. The RSBQ’s domains include General Mood, Breathing Problems, Hand Behaviors, Repetitive Face Movements, Body Rocking/Expressionless Face, Night-time Behaviors, Fear/Anxiety, and Walking/Standing. (Notably, 38 of the 45 items fall into those subscales, while 7 items remain uncategorized but still count toward the total score.) A higher RSBQ score indicates more severe or frequent Rett-related behavioral symptoms. When it was first validated, the RSBQ showed good internal consistency and test–retest reliability, confirming that it reliably captures the behavioral phenotype of RTT across different observers and over time.

Since its introduction, the RSBQ (British English version) has become one of the most widely used outcome measures in Rett Syndrome clinical studies. It is valued for its specificity to RTT’s hallmark behaviors, which general developmental scales often miss. In fact, the RSBQ has been accepted by the U.S. FDA as a primary efficacy endpoint in recent RTT drug trials. For example, the pivotal trial of trofinetide (the first FDA-approved treatment for RTT) used improvement in RSBQ scores as a co-primary outcome to demonstrate drug efficacy.^5^ The widespread adoption of the RSBQ in research and therapeutics development attests to its importance. However, until now this instrument has been available and validated only in British English, which poses a barrier in non-English-speaking regions. Many clinicians and families in Hispanic regions have had to use the RSBQ informally or rely on ad-hoc translations, limiting their ability to systematically track symptoms or participate in international studies. Given the increasing number of RTT diagnoses in Spanish-speaking countries, there is a clear imperative to produce an official Spanish translation of the RSBQ. This would allow caregivers and health professionals to communicate Rett symptoms in their native language and obtain standardized scores, just as their English-speaking counterparts can.

### Objectives

In light of the rising RTT diagnoses in Latin America and the push for inclusive research tools, the primary objective of the current initiative is to produce an accurate, culturally adapted Spanish language version of the Rett Syndrome Behaviour Questionnaire. By doing so, we aim to enable Spanish-speaking caregivers and health providers to better characterize Rett symptoms using a standardized instrument, thereby improving clinical care and research outcomes. A validated Spanish translation of the RSBQ will empower families to participate in research and therapeutic programs without language barriers. As one caregiver from Colombia expressed, involving local Rett families in this translation effort allows them “to participate in the establishment of therapeutic programs across Latin America”. In sum, having the RSBQ available in Spanish is a crucial step in globalizing Rett Syndrome efforts – ensuring that no population is left behind in the understanding and treatment of this rare disorder. The collaboration between industry researchers and RTT advocacy groups in this project exemplifies the convergence of patient-centric focus and scientific innovation, paving the way for more equitable rare disease research in the region.

## METHODS

### Ethics committee study approval

The Rett Syndrome Behaviour Questionnaire (RSBQ) study involved voluntary caregiver participation through virtual interviews and online surveys. The Institutional Review Board (IRB) at the University of Massachusetts Boston reviewed and approved the study protocol (IRB ID #3700).

### Phase 0: Spanish Language Translation and Clinical Review

The original English language RSBQ was translated into Spanish by a certified medical translator based in Colombia. The resulting translation was back-translated by a native English- and Spanish-speaking clinician and both versions were reviewed by a pediatric neurologist with native Spanish and fluent English ability.

### Phase I: Guided Administration and Cognitive Debriefing

During Phase I, native Spanish-speaking RTT caregivers were recruited through direct outreach via email, telephone, and caregiver and patient advocacy network referrals. Once contact was established, each caregiver was scheduled for a virtual one-on-one session with the investigator. This synchronous format allowed for real-time verification of comprehension and provided caregivers with a supportive environment in which to complete the questionnaire.

During each session, the investigator read each RSBQ item aloud one question at a time, allowing caregivers to respond without haste. Importantly, no responses were influenced in any way. Clarifications were provided only when a participant indicated that a question or descriptor was unclear. This approach maximized accuracy in item interpretation, reduced linguistic misunderstanding and bias, and preserved caregiver autonomy. This stage functioned as a cognitive debriefing process essential for linguistic validation, allowing the research team to observe how Spanish-speaking caregivers naturally understood each item and whether cultural or linguistic adjustments were required.

Participants who requested additional clarification or made unprompted observations about the questions were further asked for suggested modifications to the language used in the question. Responses were compiled across all participants to identify questions with higher probability of misunderstanding or request for clarification. Consensus modifications for each such question across all participants were implemented following review by two native Spanish-speaking clinicians and the rest of the research team.

### Phase II: Test-Retest Evaluation

#### Survey Timeline and Data Collection

Responses to the revised Spanish RSBQ surveys were collected between July 7, 2024, and September 20, 2025 using virtual recruitment involving Rett syndrome patient advocacy organizations across the Americas, resulting in an initial dataset of 134 caregiver submissions. Participants were recruited through a coordinated outreach effort involving Rett syndrome foundations, patient advocacy groups, and regional caregiver networks across Latin America. Leaders of these organizations played a central role in disseminating information about the study. They helped invite families to the study website, facilitated communication with caregivers in their respective countries, and ensured that the invitation reached both newly diagnosed families and long-standing members of Rett communities. This collaborative approach allowed the study to access a broad, diverse caregiver population while maintaining ethical boundaries and voluntary participation.

After one week, participants automatically received an invitation to complete a second, independent administration of the same questionnaire. This timing allowed for assessment of test–retest stability, a critical component of instrument reliability.

#### Data Cleaning and Duplicate Identification

A systematic data quality review was performed before analysis. Duplicate responses were identified by cross-referencing caregiver names, email addresses, and submission timestamps. This step revealed 15 duplicate entries representing 15 caregivers who had submitted more than once.

To maintain data integrity and avoid inflation of sample size, duplicates were removed using the following criteria:

- When multiple entries existed, the most complete and most recent submission was retained.
- All incomplete or earlier duplicate entries were removed.

After this cleaning process, the Phase II dataset was reduced from 134 to 118 unique caregiver entries, which formed the final sample for descriptive and comparative analyses.

### Statistical Analysis

Plotting and corresponding statistical analyses were performed using GraphPad Prism version 11.0.0 for Windows (GraphPad Software, Boston, Massachusetts USA). Details of statistical tests are indicated in the results. A threshold of p < 0.05 was used to determine statistical significance. Internal correlation coefficient calculation was performed using IBM SPSS Statistics for Windows, v. 31 (IBM Corp., Armonk, N.Y., USA), based on a 2-way mixed-effects model, absolute-agreement type, and mean-rating (k=2 for two repeated measurements).

## RESULTS

### Phase I Caregiver Interview

While only modest modifications were made to the certified medical translation, a number of questions were modified more significantly due to frequent requests for clarification, culturally inappropriate word choice, and other reasons. Table 1 highlights the most critical changes in wording. Importantly, while some feedback received included suggestions to add in “not applicable” or other question modifications, the original scoring and question order of the English RSBQ was left unchanged and only wording was modified in select questions. Of the 45 questions, 12 were left unchanged from the certified medical translation, 7 were critical changes causing confusion or linguistic misunderstanding (shared in Table 1), and the rest consisted of minor grammatical or word order changes. The revised questions were reviewed by the research team and finalized into the final Spanish RSBQ implemented in the online survey in Phase II.

**Table 1.** List of the questions most frequently asked for clarification by caregivers, along with the revised wording.

| Number | Original Question | Certified Translation | Modified Question | Reverse Translation | Justification |
| --- | --- | --- | --- | --- | --- |
| 4 | Makes repetitive movements involving fingers around tongue. | Realiza movimientos repetitivos que involucra el uso de los dedos alrededor de la lengua. | Realiza movimientos repetitivos alrededor de la lengua con el uso de los dedos de la mano. | Performs repetitive movements around the tongue using fingers. | Better explains the question. |
| 6 | Air or saliva is expelled from mouth with force. | Expulsa aire o saliva de la boca de manera forzosa. | Expulsa de manera forzosa, aire o saliva de su boca. | Forcefully expels air or saliva from mouth. | Improved caregiver understanding. |
| 16 | There are times when she appears miserable for no apparent reason. | Hay momentos en que parece miserable sin alguna razón aparente. | Hay momentos en que aparenta cabizbaja o triste sin alguna razón aparente. | There are moments when she appears downcast or sad, without any apparent reason. | Frequently asked for clarification because "miserable" has different, mostly negative, connotations depending on the region. |
| 20 | Hand movements are uniform and monotonous. | Los movimientos de las manos son uniformes y monótonos. | Los movimientos de sus manos tienen un patrón invariable y repetitivo. | Hand movements follow an invariable and repetitive pattern. | The use of the words "uniforme" and "monótono" was unclear to many caregivers. |
| 23 | Although can stand independently tends to lean on objects or people. | Aunque puede pararse de forma independiente, tiende a apoyarse sobre objetos o personas. | En caso de que pueda pararse de manera independiente, tiende a buscar apoyo sobre objetos o personas. | In the event that they are able to stand independently, they tend to seek support from objects or people. | Frequently asked for clarification for patients unable to stand or walk. |
| 36 | Vocalises for no apparent reason. | Vocaliza sin razón aparente. | Vocaliza o articula sonidos sin razón aparente. | Vocalizes or makes sounds without apparent reason. | Frequently asked for clarification regarding "vocalizar" due to multiple connotations in Spanish. |
| 39 | Walks with stiff legs. | Camina con las piernas rígidas. | En caso que aplique, camina con las piernas rígidas. | If applicable, walk with stiff legs. | Frequently asked for clarification for patients unable to stand or walk. |

### Participation Flow and Dataset Integrity

A total of 134 survey responses were initially collected during Phase II between July 7, 2024, and September 21, 2025. After merging entries and conducting a row-by-row verification, 15 duplicate submissions were identified, accounting for 15 caregivers who had submitted more than once. Duplicates were detected by cross-referencing caregiver name, email address, and submission timestamps. In each case, the most recent and most complete submission was retained, and earlier duplicated entries were removed.

### Demographic and Geographic Distribution

Caregivers represented a wide range of countries across Latin America (Table 2), reflecting the success of the recruitment process coordinated through Rett organizations and advocacy leaders. The vast majority of participants (47/51) were female caregivers, consistent with global caregiving trends in Rett Syndrome.^6,7^

**Table 2.** Geographic distribution of the final 51 study participants who completed test and re-test surveys and passed all quality control checks.

| **Country** | **Number of Responders (%)** |
| --- | --- |
| **Argentina** | 20 (39.2%) |
| **Mexico** | 16 (31.4%) |
| **Peru** | 6 (11.8%) |
| **Colombia** | 3 (5.9%) |
| **Uruguay** | 3 (5.9%) |
| **Guatemala** | 2 (3.9%) |
| **United States (Spanish-speaking caregiver)** | 1 (2.0%) |

This distribution highlights the robust reach of the Spanish RSBQ validation effort across Latin America and demonstrates meaningful engagement from both South American and Central American caregiver populations. The presence of a small number of participants outside the region further underscores the broad relevance of the translated instrument for Spanish-speaking families globally.

### Phase II Test–Retest

A total of 51 unique caregivers completed both timepoints and met all quality control criteria. RSBQ total scores ranged from 17 to 90, representing a diverse range of patient severities with high retest correlation (Fig. 1). A Bland-Altman plot of the paired responses indicated 92.2% (47/51) response averages were within 2 standard deviations of the difference (Fig 2A). Responses were linearly correlated for each participant (Fig. 2B). RSBQ total score test-retest intraclass correlation coefficient (ICC) estimate with 95% confidence intervals (CI) for single measures was 0.907 (95% CI, .843 to .946).

**Figure 1.**
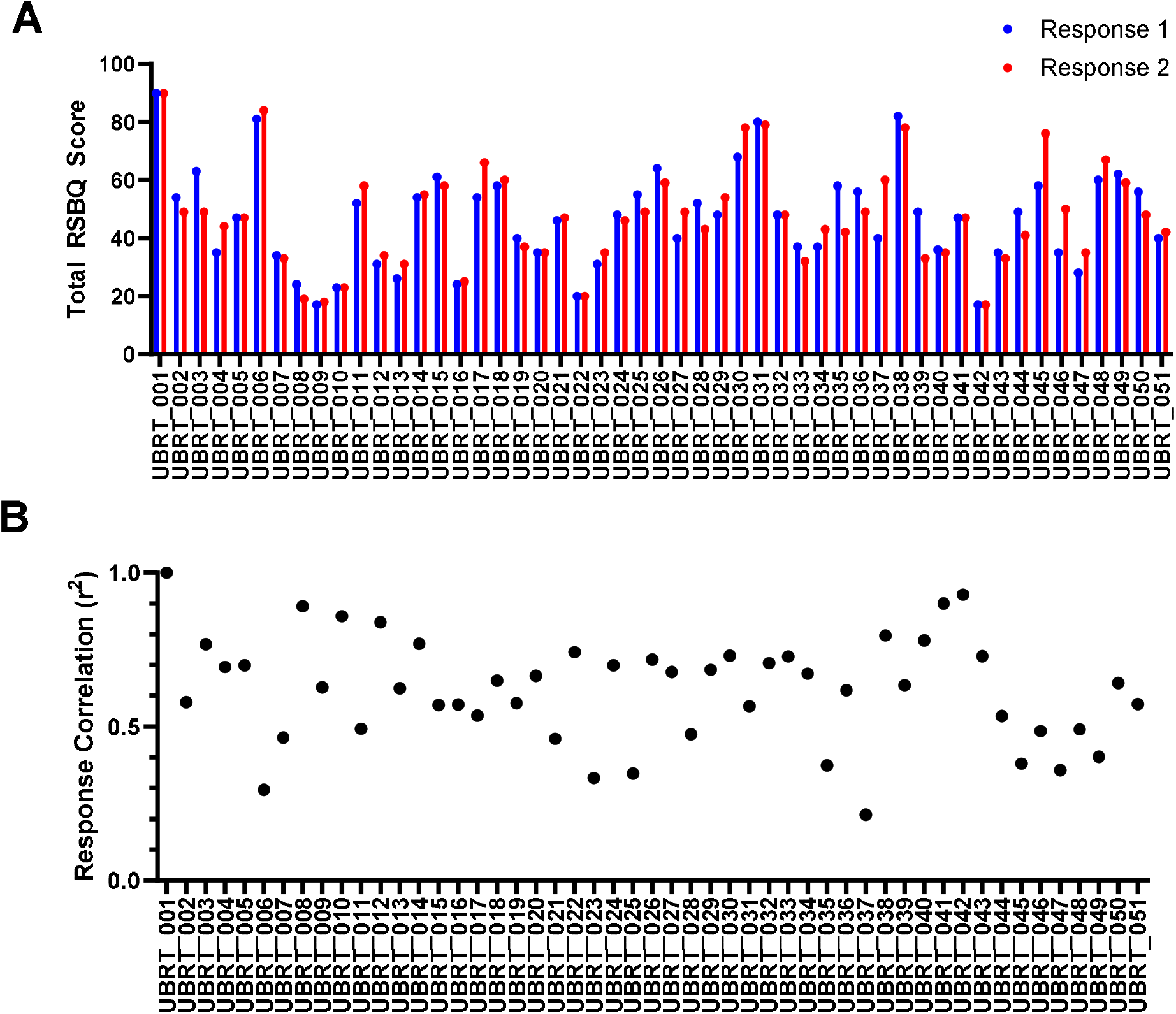
(A) RSBQ total scores are shown for each study participant at the test (Response 1) and retest (Response 2) timepoints. (B) Pearson correlation coefficient for each participant shows overall high correlation of responses between the test and retest.

**Figure 2.**
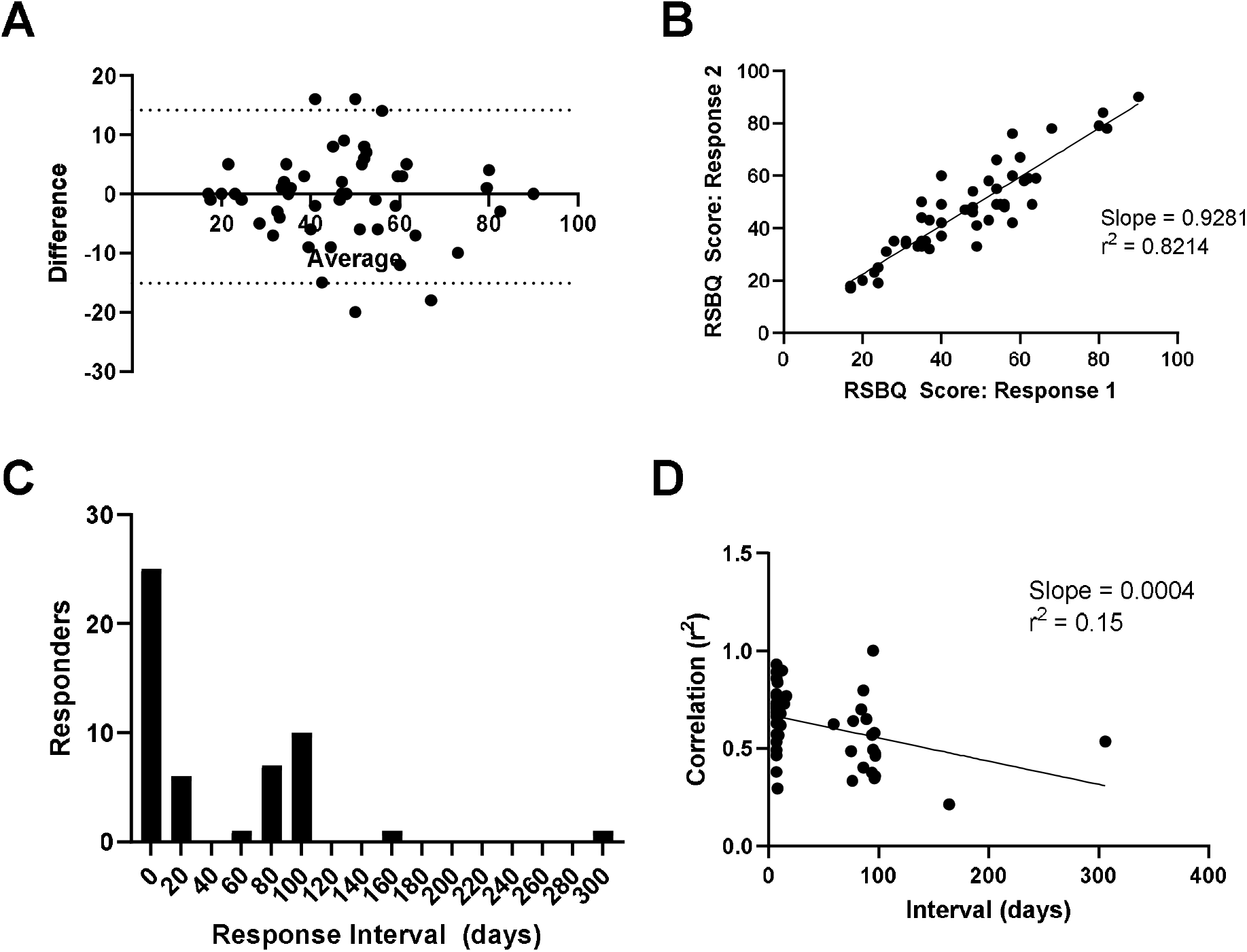
(A) Bland-Altman plot of response score difference vs. mean shows 47 out of 51 responders landing within two standard deviations. (B) The test and retest responses were significantly linearly correlated (slope = 0.9281, Pearson r^2^ = 0.8214, F test non-zero slope p < 0.0001). (C) Histogram of the response intervals shows multimodal responses with a majority of responses occurring within the expected 1-2 weeks. (D) A slight but statistically significant effect of time on the correlation coefficient for each responder was observed (slope = 0.0004, Pearson r^2^ = 0.15, F test non-zero slope p < 0.005).

Additional reminders and community events were implemented by the regional organizations to reach the target goal of 50 study participants. This resulted in some responders having far greater intervals between the test and retest than initially planned. The mean response interval was 45.4 days, a median of 11 days, and a standard deviation of 56.5 days. The distribution of responses clustered into two main groups of 7-8 days and 80-100 days (Fig. 2C). No significant mean difference in RSBQ scores were observed between the two timepoints (Test 1: 46.8 +/-17.0; Test 2: 47.2 +/-17.4; paired Student’s t-test p = 0.6540). A slight but statistically significant correlation of interval between responses on the Pearson correlation coefficient for each participant was observed (Fig. 2D).

Due to the natural progression of RTT patients, it is possible that differences in scoring for longer inter-test periods may reflect real changes in patient symptoms, which could confound test-retest analysis. To remove this potential confounding factor, a secondary analysis was conducted on data restricted to participants who responded within 1 and 3 weeks (7-21 days) between the two test administrations (Fig. 3A). This time range corresponded to the main cluster of 31 responders (Fig. 2C). A Bland-Altman plot of the paired responses indicated 90.3% (28/31) response averages were within 2 standard deviations of the difference (Fig 3B). Responses were linearly correlated for each participant (Fig. 3C). To evaluate whether specific questions may have greater intra-rater variability between the test and retest, we evaluated the mean difference between timepoints for each question individually (Fig. 3D). Kruskal-Wallis testing did not indicate a significant effect of the question on the mean difference (p = 0.7392). This suggests that the translations, including the specific questions with adjusted wording, yielded consistent responses between the test and retest timepoints.

**Figure 3.**
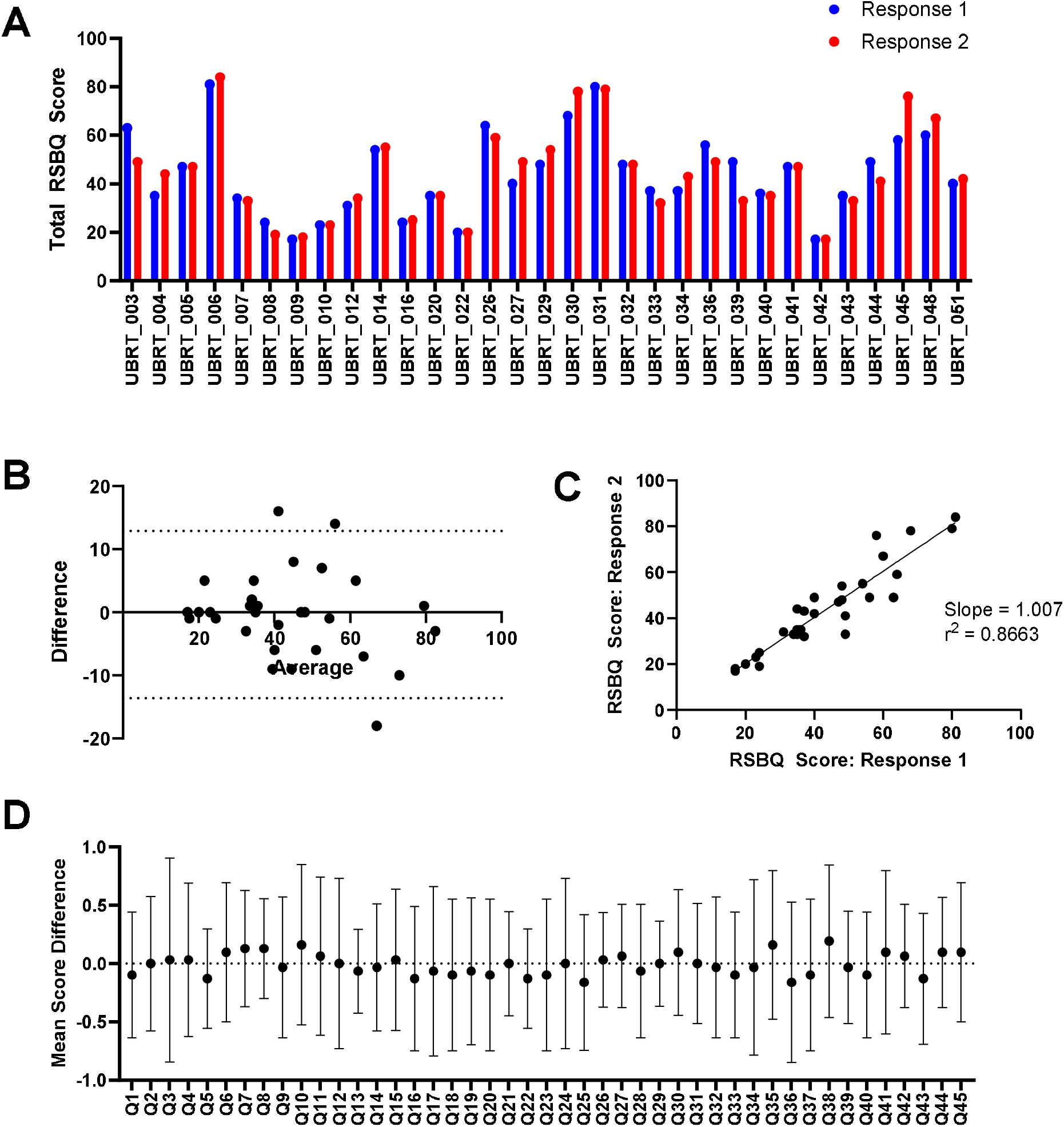
(A) RSBQ total scores are shown for 31 study participants completing the test-retest surveys within 21 days at the test (Response 1) and retest (Response 2) timepoints. (B) Bland-Altman plot of response score difference vs. mean shows 28 out of 31 responders landing within two standard deviations. (C) The test and retest responses were significantly linearly correlated (slope = 1.007, Pearson r^2^ = 0.8663, F test non-zero slope p < 0.0001). (D) Score differences of responses for all 31 participants show a consistent pattern with the mean centered at 0 and consistent variance. Kruskal-Wallis testing did not indicate a significant effect of the question on the mean difference (p = 0.7392). Error bars indicate standard deviation.

## DISCUSSION

The present study represents the first structured effort to validate a Spanish translation of the Rett Syndrome Behaviour Questionnaire (RSBQ) through direct engagement with Spanish-speaking caregivers across Latin America. This work addresses a critical gap in the availability of standardized behavioral assessment tools for RTT in non-English-speaking populations. As the incidence and recognition of Rett syndrome steadily increases in Latin America, the need for accessible, culturally appropriate clinical instruments has become more urgent.

### Importance of a Spanish RSBQ in Emerging RTT Contexts

Historically, behavioral and clinical research in RTT has relied predominantly on English-language tools, which limits the participation of families and clinicians in regions such as Latin America. The RSBQ — widely used in clinical trials and approved by regulatory bodies as a key efficacy endpoint — plays a central role in both routine clinical evaluation and therapeutic decision-making. However, without a validated Spanish version, clinicians and researchers in Spanish-speaking countries have lacked a consistent method to capture the severity and progression of RTT symptoms.

The demographic distribution observed in this study underscores this urgency: the majority of participants were from Argentina, Mexico, and Peru — countries where RTT is increasingly recognized, and where caregiver organizations are expanding their advocacy and diagnostic support networks. The strong participation from these regions reflects both the clinical need and the readiness of Latin American families to contribute to RTT research when barriers such as language are addressed.

### Caregiver Engagement and Feasibility of the Validation Approach

The two-phase design — combining guided administration with an independent test and retest— proved both feasible and effective for linguistic validation in a geographically dispersed population. Caregivers demonstrated high engagement during Phase I’s guided sessions, which allowed investigators to directly observe linguistic challenges, clarify ambiguous wording, and ensure accurate item comprehension without influencing responses. This provided a rich qualitative foundation for confirming conceptual equivalence between the Spanish and English versions of the instrument.

The Phase II retest further demonstrated feasibility of completing both survey administrations. The support of regional patient advocacy organizations was instrumental in retaining participants through their respective communication channels that highlighted the importance of participation in supporting the clinical care and research capabilities for the RTT communities in their respective countries. These findings suggest that Spanish-speaking caregivers are willing and able to participate in multi-step research processes, particularly when supported by advocacy organizations and when the study holds direct relevance to clinical care.

### Implications for Clinical Research and Healthcare Systems in Latin America

The findings from this study extend beyond linguistic validation. They reveal a highly mobilized caregiver community, strong participation from Latin American RTT foundations, and a growing regional infrastructure capable of supporting clinical research. Several hospitals and regional clinical centers have already expressed interest in integrating the Spanish RSBQ into their diagnostic pathways. This reflects the tool’s potential to:

- Improve early recognition of RTT symptoms,
- Standardize behavioral assessments across Spanish-speaking regions,
- Reduce the cost and burden associated with complex diagnostic evaluations, and
- Support eligibility screening and endpoint assessment for upcoming therapeutics.

As countries in Latin America increase their diagnostic capability and focus on rare diseases— including recent legislative advances in nations such as Panama—validated, linguistically appropriate clinical instruments will be essential for integrating RTT into public health systems.

### Strengths and Limitations

A major strength of this study is its direct caregiver involvement and broad geographic reach across Latin America. The guided administration method ensured high comprehension of the translated items and identification of potentially confusing question wording while enabling participants to propose alternative language to improve comprehension and linguistically appropriate context. The test-retest phase provided a strong foundation for evaluating stability and reliability across a larger number of participants spanning a wide geographic range. Engagement from multiple RTT foundations also strengthened recruitment and allowed caregivers from diverse socioeconomic and cultural backgrounds to participate.

However, several limitations must be acknowledged. First, although all participants were Spanish-speaking, regional linguistic variation (“regionalisms”) across Latin America introduced natural differences in vocabulary, phrasing, and interpretation. These differences required careful clarification during guided sessions and must be continuously considered in future iterations of the instrument, especially for countries with highly distinct dialects.

Second, participant time constraints presented challenges. Many caregivers reported limited availability due to work demands, significant caregiving responsibilities, and household duties. RTT caregivers often manage complex medical routines, which may have impacted completion times and Phase II follow-up rates. These constraints are inherent to real-world caregiver populations but should be recognized as potential contributors to variability in participation.

Third, technology familiarity varied considerably across participants. While some caregivers navigated digital forms easily, others required assistance due to limited experience with online platforms, unstable internet connections, or device limitations. These differences may have influenced their ability to complete surveys promptly or independently during Phase II.

Fourth, the study relied on voluntary participation through advocacy networks, which may introduce selection bias toward families who are more engaged, better connected, or more technologically equipped. This could lead to underrepresentation of rural or lower-income caregivers who face barriers to digital participation. Included in this limitation is the geographic bias of the results. Notably absent are participants from the Caribbean region as well as Spain.

### Overall Significance

Taken together, the results demonstrate that the Spanish RSBQ is not only feasible and acceptable to caregivers but also urgently needed to support clinical care and research readiness in Latin America. This study lays the foundation for a validated Spanish behavioral instrument that can be incorporated into routine clinical evaluation, early diagnosis efforts, and multinational clinical trials.

The linguistic validation of a Spanish-language translation of the Rett Syndrome Behaviour Questionnaire (RSBQ) represents a significant advancement for clinical care, research accessibility, and health equity across Latin America. By providing caregivers and clinicians with a culturally appropriate instrument, this work directly strengthens caregiver participation in diagnostic pathways and clinical decision-making and expands opportunities for participation in global RTT research initiatives. A validated Spanish RSBQ has immediate implications for improving caregiver engagement in the diagnosis and care of RTT patients in Spanish-speaking populations. Historically, delayed diagnosis has been common in Latin America due to limited access to specialized neurological services, under-recognition of RTT symptoms, and the absence of standardized behavioral assessment tools in the region. With the availability of a linguistically-validated Spanish RSBQ:

- Clinicians can more reliably identify core RTT behaviors
- Caregivers can communicate symptom progression with greater clarity
- Diagnostic uncertainty can be reduced through consistent, structured behavioral reporting

As several hospitals and regional RTT centers have already expressed interest in adopting the Spanish RSBQ, its integration into clinical workflows may reduce the burden of diagnostic evaluations, optimize resource allocation, and support earlier referral to specialized services.

### Research Readiness and Global Trial Inclusion

The growing interest in RTT research in Latin America underscores the importance of standardized, validated outcome measures for Spanish-speaking participants. The availability of this instrument supports RTT behavioral assessment for research, eligibility screening for clinical trials, cross-cultural comparability of RTT severity scores, and meaningful integration of Latin American populations into international datasets. This Spanish RSBQ positions Latin America as an increasingly relevant region for rare disease research and strengthens the infrastructure needed for multinational clinical trials.

### Empowerment of Caregivers and Advocacy Networks

The high level of caregiver participation in this study highlights the strong commitment of RTT families in Latin America. The guided approach of Phase I not only produced high-quality data but also strengthened caregiver trust and engagement, reinforcing the role of families as central partners in rare disease research. Furthermore, the collaboration with RTT foundations across multiple countries demonstrates the power of community-led mobilization and the critical role of patient advocacy in bridging gaps between scientific innovation and real-world clinical needs. As these organizations continue to expand outreach, the Spanish RSBQ can become a unifying resource for awareness, education, and long-term caregiver support.

### Future Directions

Although this phase included extensive representation from Mexico, Peru, Argentina, and other Latin American countries, future work may expand to Spain, the Caribbean, and underrepresented populations to ensure broader linguistic neutrality and regional sensitivity. The methodology used here — guided comprehension testing, follow-up retesting, and multi-country recruitment — can serve as a model for adapting other clinical instruments into Spanish. As companies advance new RTT therapies in Latin America, a linguistically validated Spanish RSBQ will be essential for outcome measurement, regulatory submission, and post-market monitoring in Spanish-speaking cohorts.

### Overall Impact

This study marks a foundational contribution to improving Rett Syndrome caregiver participation and research inclusivity in the Spanish-speaking world. By creating the first structured Spanish-language adaptation of the RSBQ, this project advances clinical equity, empowers caregivers, and strengthens the infrastructure needed for multinational therapeutic development. The Spanish RSBQ is poised to become a critical tool for clinicians, researchers, and families across Latin America and beyond.

## Data Availability

All data produced in the present work are contained in the manuscript. The translated scale is available upon reasonable request to the authors.

## ACKNOWLEDGMENTS

The authors are grateful to Dr. Anthony Charman for permission to translate the English RSBQ into Spanish and to the foundations and organizations involved in participant recruitment and study awareness across the Spanish-speaking RTT community. We would like to specifically acknowledge Paula Ferreira Goldke (Associação Brasileira de Síndrome de Rett (Abre-te), Brazil); Clarivel del Castillo Barrientos (Asociación Síndrome de Rett de Guatemala); Jessica Denise Cubillos Arellano (Fundación Caminamos por Ellas y Ellos – Rett Syndrome, Chile); Marcia Elizabeth Cadena (Fundación Rett Ecuador, Ecuador); Fabián Alberto Crespo (Fundación Síndrome de Rett, Argentina); Fabiana Pinedo Cruz (Asociación Peruana de Síndrome de Rett – Miradas que hablan, Peru); Miriam Salinas Aguirre (Asociación Rettos con el Corazón A.C. de C.V., Mexico); and Daniela Lima, (Rett Syndrome Support Group of Uruguay), and the International Rett Syndrome Foundation, USA. Their collective efforts in advocacy, family support, coordination, and community building made it possible to ensure that this project meaningfully reflects the voices and needs of families affected by Rett syndrome in Latin America. This work would not have been possible without their generosity, collaboration, and shared commitment to advancing research and care in the region. The study was funded by Unravel Biosciences, Inc.

